# Infections in childhood and subsequent diagnosis of autism spectrum disorders and intellectual disability: a register-based cohort study

**DOI:** 10.1101/2021.03.10.21252968

**Authors:** Håkan Karlsson, Hugo Sjöqvist, Martin Brynge, Renee Gardner, Christina Dalman

## Abstract

**Objective:** To explore the associations between childhood infections and subsequent diagnoses of autism spectrum disorder (ASD), intellectual disability (ID) and their co-occurrence.

**Methods:** The association between specialized care for any infection, defined by ICD-codes and later ASD or ID was investigated in a register-based cohort of 556,732 individuals born 1987-2010, resident in Stockholm County, followed from birth to their 18th birthday or December 31, 2016. We considered as potential confounders children’s characteristics, family socioeconomic factors, obstetric complications, and parental histories of both treatment for infection and psychiatric disorders in survival analyses with extended Cox regression models. Residual confounding by shared familial factors was addressed in sibling analyses using within-strata estimation in Cox regression models. Sensitivity analyses with exclusion of congenital causes of ASD/ID and documented risk for infections were also performed.

**Results:** Crude estimates indicated that infections during childhood were associated with later ASD and ID with largest risks observed for diagnoses involving ID. Inclusion of covariates, exclusion of congenital causes of ASD/ID from the population and sibling comparisons highlighted the potential for confounding by both heritable and non-heritable factors, though risks remained in all adjusted models. In adjusted sibling comparisons, excluding congenital causes, infections were associated with later ‘ASD without ID’ (HR 1.24, 95%CI 1.15-1.33), ‘ASD with ID’ (1.57, 1.35-1.82) and ‘ID without ASD’ (2.01, 1.76-2.28). Risks associated with infections varied by age at exposure and by age at diagnosis of ASD/ID.

**Conclusions:** Infections during childhood cannot be excluded in the etiology of ASD, particularly ASD with co-occurring ID.

## 1. Introduction

Autism spectrum disorder (ASD) is diagnosed throughout childhood and age at diagnosis appears to be determined by both internal (severity, sex) and external factors (parental educational level, access to care) ^1^. While parents often report deviations from normal developmental trajectories at an early age, the pattern of onset varies substantially. It remains to be established whether children later diagnosed are ‘born’ with ASD ^2, 3^ or if the early childhood environment may contribute to a trajectory towards ASD in vulnerable children.

The etiology of ASD remains elusive but appears to involve both heritable and non-heritable genetic variants and environmental exposures ^4^. The relative contributions of these plausibly causal factors vary across diagnostic sub-groups ^5-7^. Intellectual disability (ID, IQ<70) is diagnosed in around 1/3 of those diagnosed with ASD ^8^. Individuals diagnosed with ASD without co-occurring ID are more likely to have a family history of ASD and other psychiatric diagnoses than individuals diagnosed with ASD with ID ^9, 10^. Moreover, ID in individuals with ASD appears less familial compared to ID in the absence of ASD and may therefore have non-heritable causes ^11^.

Infections that can invade the fetal brain are established causes of life-long behavioral and intellectual disabilities ^12, 13^. Infections targeting the brain during childhood have also been associated with cognitive impairments and psychiatric illness which become evident only later in life ^14-18^. A growing literature associates childhood infections with no apparent CNS involvement with schizophrenia risk in adulthood ^19-22^, with a significant portion of risk mediated by reduced cognitive abilities ^21^. Observations of deficits in innate immunity among neonates who later develop schizophrenia ^23^ or ASD ^24^ further highlight the potential contribution of postnatal infections to these outcomes ^25-27^. However, only a handful of studies, with equivocal results, have investigated the relationship between childhood infections and the risk of ASD ^18, 28-31^.

We aimed to explore the associations between infections and subsequent diagnoses of ASD, ID and their co-occurrence, and to understand whether any observed associations varied by age at infection.

## 2. Methods

### 2.1 Study population and general design

We defined a register-based cohort of 556,732 individuals born 1987-2010, resident in Stockholm County for ≥3 years ^32^. Individuals not born in Sweden, adopted, not registered in the Medical Birth Register (MBR), or missing maternal data were excluded from the study (Figure S1).

Exposure, outcome and covariate data were extracted from national and regional registers containing routinely collected health and sociodemographic data cross-linked via each resident’s unique national identification number ^33^. The study was approved by the Stockholm regional ethics review board (DNR 2010/1185-31/5). Informed consent was not required to use anonymized register data.

### 2.2 Diagnoses of ASD and ID

Diagnostic outcomes as of December 31, 2016 were defined by validated procedures covering all inpatient and outpatient pathways to care and diagnosis in Stockholm County ^11, 32^ (ICD-9: ASD 299, ID 317-19, ICD-10: ASD F84, ID F70-79). We considered two overlapping diagnostic groups (individuals who received any diagnosis of ASD or any diagnosis of ID), as well as the three mutually exclusive diagnostic outcomes: ASD with co-occurring ID (‘ASD with ID’), ASD without co-occurring ID (‘ASD without ID’) and ID without co-occurring ASD (‘ID without ASD’), Figure 1A.

**Figure 1.**
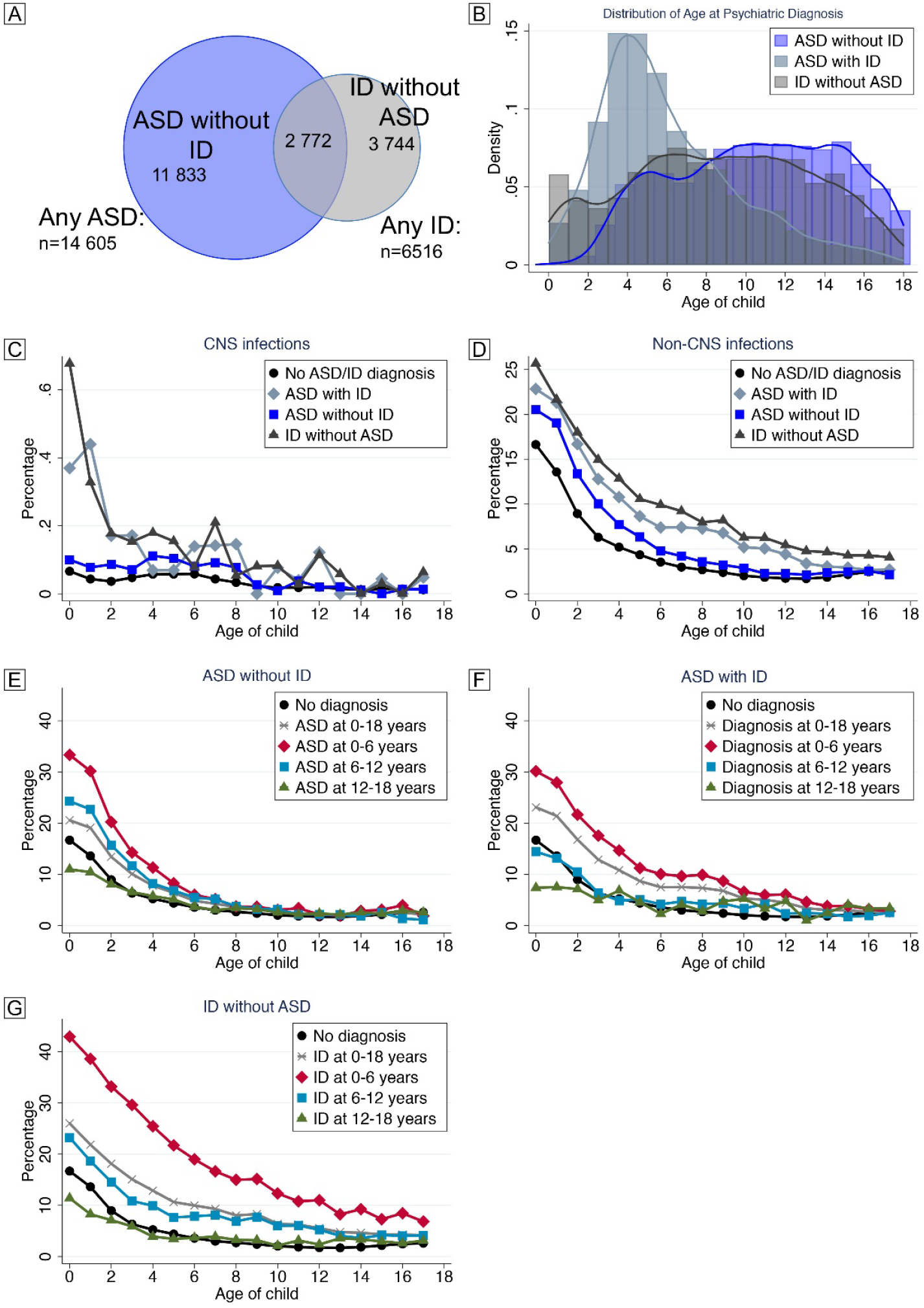
Description of outcomes and exposures in the study population. Individuals diagnosed with ASD and/or ID identified in the final study population (n=556,732) as of December 31, 2016 (A). Age at first diagnosis of ASD/ID for the mutually exclusive diagnostic groups (B). Incidence of CNS (C) and non-CNS infections (D) according to age among children unaffected by ASD/ID and among those diagnosed with ‘ASD without ID’, ‘ASD with ID’, ‘ID without ASD’. Note the different scales of the y-axes in C and D. Incidence of one or more infections according to age among children unaffected by ASD/ID and those diagnosed with ‘ASD without ID’ (E), ‘ASD with ID’ (F) and ‘ID without ASD’ (G), throughout childhood and stratified by age at diagnosis of ASD/ID.

### 2.3 Assessment of childhood infections

All in- or outpatient specialized care diagnoses of infection, defined by ICD-9/10 codes as detailed previously ^19^, were identified in the National Patient Register (NPR). We omitted all codes “sequel” and “post.”

### 2.4 Covariates

Covariates whose distribution varied by exposure to infection between birth and age 18 (Table 1) and that were associated with ASD or ID (Tables S1 & S2) were considered as potential confounders. Parental region of origin was identified from the Multigenerational Register. The Longitudinal Integrated Database for Health Insurance and Labor Markets Studies (LISA) supplied quintiles of family income adjusted by inflation and size of family (recorded at the birth of the index person) and highest parental education. MBR provided information on maternal body mass index (BMI, categorized according to WHO standards ^34^), birth order, parental age at birth, birth season, maternal pre-eclampsia during pregnancy, size for gestational age (GA) at birth, Apgar score at 5 min after birth, birth by cesarean section, and GA at birth. NPR provided information on parental history of infections before birth of the index person. NPR and regional registers provided information on history of parental psychiatric illness (defined as ICD10 F-chapter diagnosis, ICD9 290-315, or ICD8 290-315).

### 2.5 Statistical analyses

We employed survival analysis with the extended Cox regression model using within-model stratification to account for differences by the children’s year of birth. Age of the child was used as the underlying timescale, beginning at birth. Individuals were followed until death, emigration from Stockholm, ASD/ID diagnosis, or end of study. The infection variables were treated as time-dependent exposures, in which the individual could have had an infection from birth up to 18 years of follow-up, given that the child was not diagnosed with an outcome of interest or otherwise censored. To account for the possible non-proportional risk of the outcomes over follow-up time, we considered both cumulative time wherein the exposure to infection could occur from birth up to the 18^th^ birthday, and stratified according to age at infection; 0-<1 year, 1-<3, 3-<6, 6-<12, and 12-<18. To test for differences in risk related to age at infection, we used the Wald test to evaluate the beta coefficients for each strata of age at exposure in the cumulative models. To assess if the associations varied by age at outcome, we also stratified the outcome according to age at first diagnosis of ASD and/or ID: 0-<6, 6-<12 and 12-<18 years.

To explore potential confounding by familial factors shared between siblings not accounted for by adjustment for covariates, we performed sibling analyses using within-strata estimation in Cox regression models, adjusted for sex, parity, birth season, GA at birth and cesarean section in which we discarded all only-child observations (n=131,335). Data management was done in SAS 9.4, data analyses in Stata/MP 15.1.

### 2.6 Sensitivity analyses

Individuals with genetic disorders are at increased risk for both ASD/ID and repeated specialized care for infections during infancy and childhood ^35, 36^. To address if such individuals were driving potential associations observed, we repeated the population and sibling comparison analyses after excluding individuals affected by a study outcome who were also diagnosed with a congenital disorder, as detailed in ^11^.

## 3. Results

### 3.1 Description of the study population

Of the 556,732 children included in our study (Figure S1), 282,595 (50.8%) had at least one registered infection before age 18. All investigated covariates differed significantly (p<0.001) between those who received specialized care for infection and those who did not (Table 1). Among those who were not diagnosed with an infection during childhood, 5,803 (2.1%) were diagnosed with ASD and 2,051 (0.7%) with ID. Among those diagnosed with one or more infection, 8,802 (3.1%) were diagnosed with ASD and 4,465 (1.6%) with ID (Table 1). As compared to ‘ASD without ID’ or ‘ID without ASD’, ‘ASD with ID’ was diagnosed at an earlier age (Figure 1B). CNS infections were rare as compared to non-CNS infections. Both types of infections were overrepresented in individuals with the investigated outcomes, particularly those involving ID (Figure 1C-D). Infections were more common during the first few years of childhood in all diagnostic groups (Figure 1E-G), particularly among those with an early diagnosis of ‘ID without ASD’ or ‘ASD with ID’, as compared to those without a diagnosis of ASD/ID. Infections were more common throughout childhood among children diagnosed with ID before age 12 as compared to those unaffected by ASD/ID or those diagnosed with ‘ASD without ID’.

All included covariates were associated with ASD and age at ASD diagnosis. For example, ASD was more common among males and first-born children, particularly among those diagnosed before age 6 (Table S1). Similarly, obstetric complications, such as pre-term birth and cesarean section were associated with early diagnosis whereas parental psychiatric history was more common among cases diagnosed after age 12. Similar observations were made for ID (Table S2).

### 3.2 Main analyses

#### 3.2.1 Overlapping diagnostic outcomes

Inclusion of covariates in our fully adjusted models (Figure 2) attenuated risk estimates compared to crude models (Figure S2) but did not change the overall findings. In fully adjusted models, infections during childhood (0-<18 years) were associated with later ASD (HR 1.23, 95%CI 1.19-1.28) and ID diagnoses (HR 2.16, 95%CI 2.05-2.28). Infections at all age intervals except 6-<12 were associated with later ASD, with the strongest association observed for infections in the age interval 1-<3 (HR 1.26, 95%CI 1.21-1.31) Figure 2A. Infections at all ages were associated with a later ID diagnosis, with the strongest association observed for infections during age 1-<3 (HR 2.12, 95%CI 2.00-2.24), Figure 2E.

**Figure 2.**
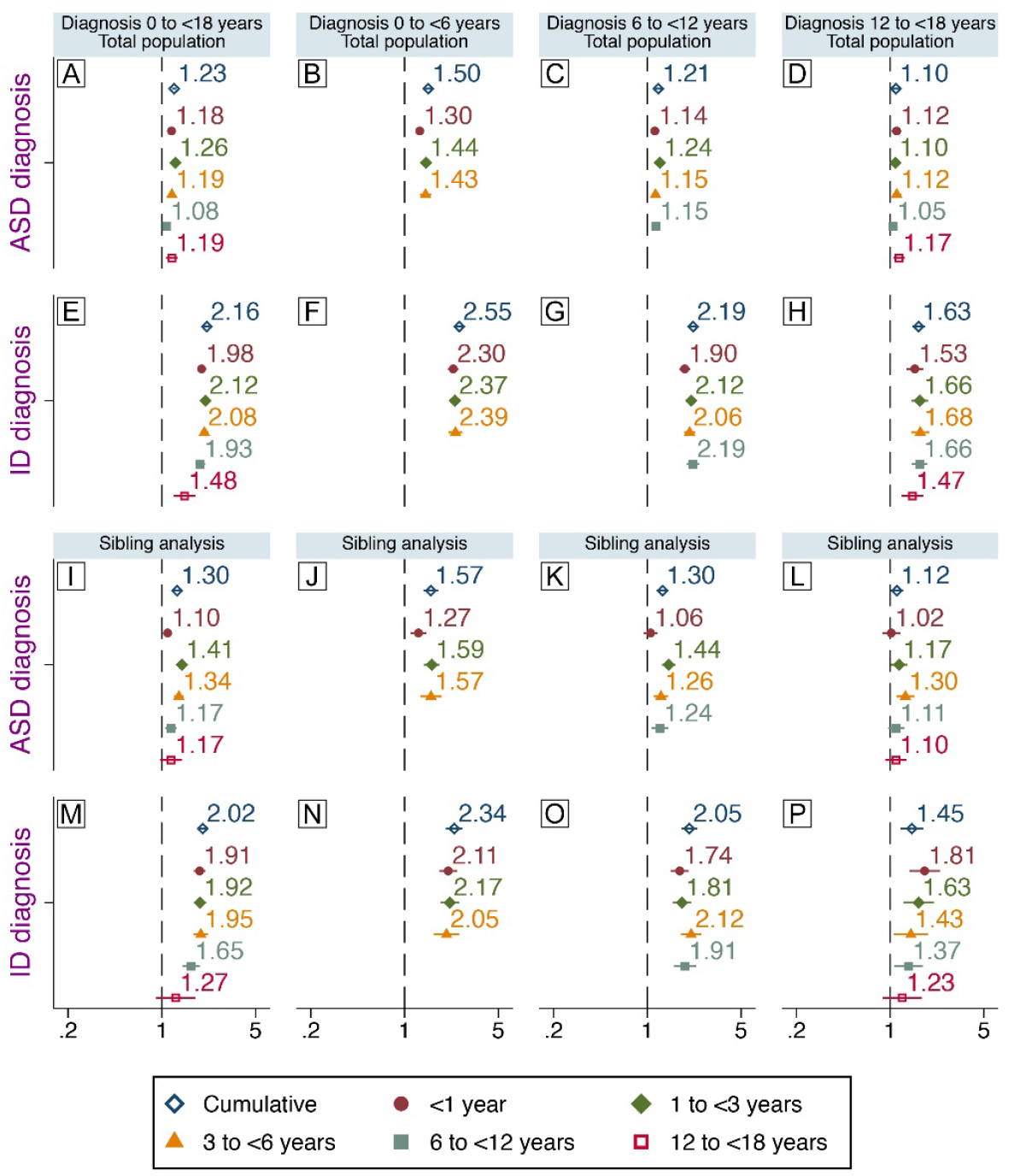
Infections during childhood and later diagnosis of ASD or ID. Associations between specialized care for infections during childhood and later, non-mutually exclusive, diagnosis of ASD and ID. Associations between exposure at different age intervals and diagnoses at different age intervals are also shown. Comparison between unrelated individuals in the general population are shown (A-H) and comparisons between full biological siblings (I-P). Hazard ratios presented here are from fully adjusted models. Population-based estimates (A-H) are adjusted for sex, parity, maternal body mass index, pre-eclampsia, parental age, education, income, region of origin, histories of psychiatric illness and infections, season of birth, gestational age at birth, size for gestational age, cesarean section, Apgar score. Estimates from the sibling analyses (I-P) are adjusted for sex, parity, gestational age at birth and cesarean section.

Infections were associated with ASD diagnosed at age 0-<6 (HR 1.50, 95%CI 1.40-1.61, Figure 2B) and 6-<12 (HR 1.21, 95%CI 1.14-1.28, Figure 2C), but not 12-<18 (Figure 2D). Infections were associated with ID diagnosed throughout the follow-up, Figures 2F-H, with the strongest association during ages 0-<6 (HR 2.55, 95%CI 2.33-2.80, Figure 2F).

Sibling comparison models resulted in generally comparable associations to the total population analysis, see Figure 2I-P. Notably, infections during age 1-<3 exhibited a stronger association with ASD diagnosed before age 12 (Figure 2J-K). Only infections 6-<12 were associated with ASD diagnosed at 12-<18 (Figure 2L). Infections at 12-<18 were not associated with ASD or ID.

#### 3.2.2 Mutually exclusive outcomes

Similar results regarding the mutually exclusive outcome groups were observed in crude (Figure S3) and adjusted models (Figure 3). Infections during childhood were associated with the mutually exclusive diagnoses with the largest risk estimates observed for ‘ID without ASD’ (HR 2.56, 95%CI 2.39-2.75, Figure 3I) and the smallest for ‘ASD without ID’ (HR 1.15, 95%CI 1.10-1.19, Figure 3A). Whereas infections at all ages were associated with ‘ID without ASD’ and ‘ASD with ID’, only those at 1-<3 and 3-<6 were significantly associated with ‘ASD without ID’. Larger point-estimates were observed among those diagnosed early. Infections were not associated with ‘ASD without ID’ diagnosed after age 12 (Figure 3B-D, F-H & J-L).

**Figure 3.**
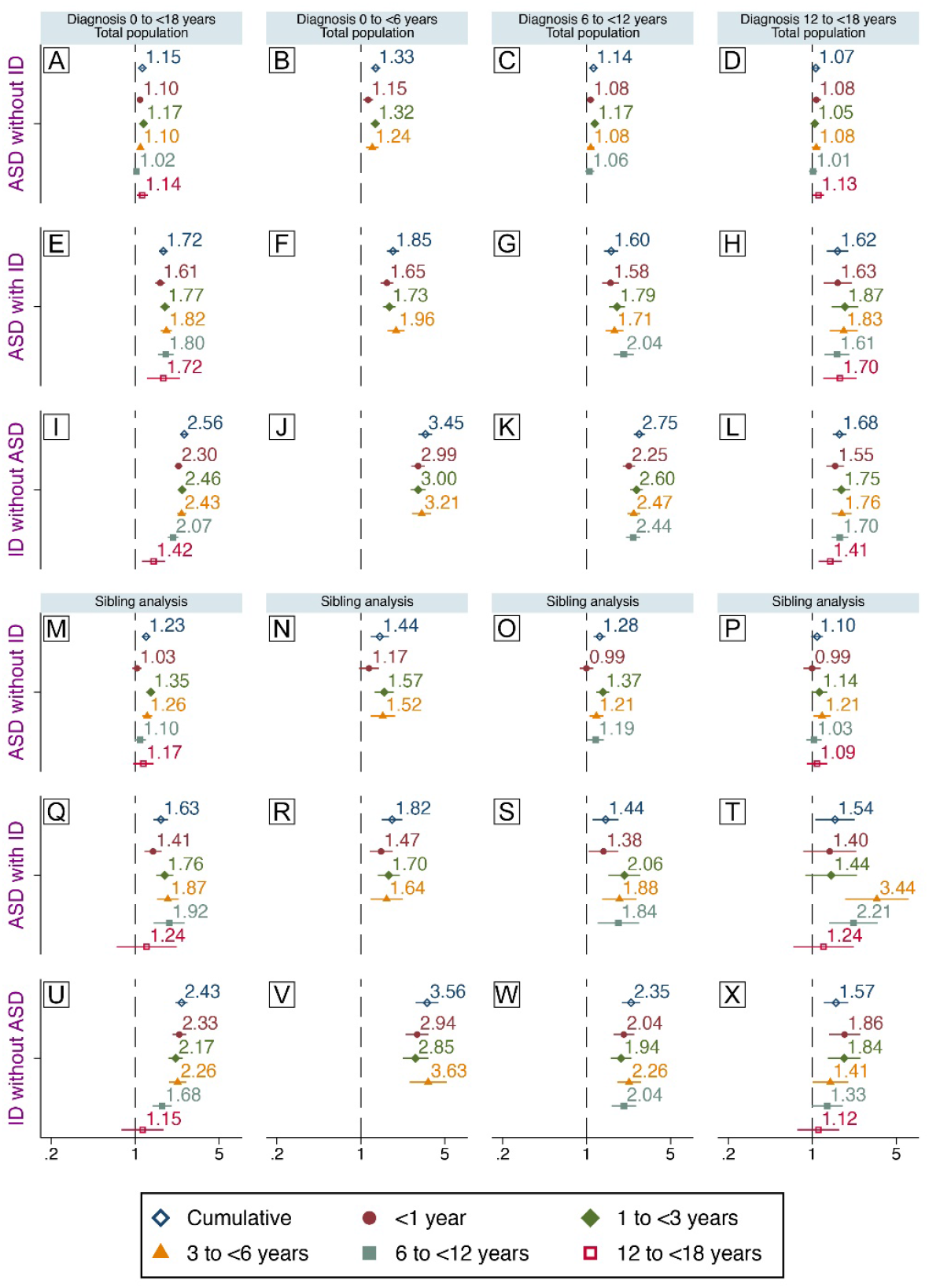
Infections during childhood and mutually exclusive diagnoses. Associations between specialized care for infections during childhood and later diagnosis of ‘ASD without ID’, ‘ASD with ID’ or ‘ID without ASD’. Associations between exposure at different age intervals and diagnoses at different age intervals are also shown. Comparisons between unrelated individuals in the general population (A-L) and between full biological siblings (M-X) are shown. Hazard ratios presented here are from fully adjusted models. Population-based estimates (A-L) are adjusted for sex, parity, maternal body mass index, pre-eclampsia, parental age, education, income, region of origin, histories of psychiatric illness and infections, season of birth, gestational age at birth, size for gestational age, cesarean section, Apgar score. Estimates from the sibling analyses (M-X) are adjusted for sex, parity, gestational age at birth and cesarean section.

Sibling analyses resulted in slightly modified associations as compared to those observed in the general population, with enhanced risk for ‘ASD without ID’ (Figure 3M-P) and mostly attenuated risks for ‘ASD with ID’ (Figure 3Q-T) or ‘ID without ASD’ (Figure 3U-Y). Associations with infections at 12-<18 were attenuated towards unity for all outcomes. Higher risk estimates for ‘ASD without ID’ were largely driven by infections in the age intervals 1-<3 and 3-<6 (Figure 3M) among those diagnosed before age 6 (Figure 3N) or at 6-<12 (Figure 3O). Infections during the first year of life were associated with later ‘ASD with ID’ (Figure 3Q) with the largest risk estimate observed among those diagnosed before age 6 (Figure 3R). Infections before age 3 were no longer associated with ‘ASD with ID’ diagnosed at 12-<18, though elevated risks were observed for infections at 3-<6 and 6-<12 (HR 3.44, 95%CI 1.87-6.33, and HR 2.21, 95%CI 1.39-3.51), respectively, Figure 3T). Infections during early childhood remained strongly associated with ‘ID without ASD’ (Figure 3U), particularly among those diagnosed before age 6 (Figure 3V) or at 6-<12 (Figure 3X), but also among those diagnosed at 12-<18 (Figure 3Y).

### 3.3 Sensitivity analyses

Exclusion of children diagnosed with congenital disorders from the total population (Figure S4) and sibling (Figure S5) comparisons attenuated the associations observed between infections and the mutually exclusive diagnoses involving ID. In the sibling comparison, infections at 1-<3 and 3-<6 remained associated with ‘ASD without ID’. Infections before age 12 remained associated with both ‘ASD with ID’ and ‘ID without ASD’ throughout the follow-up. Among those diagnosed at 12-<18, only infections before age 3 were associated with ‘ID without ASD’, whereas infections at 3-<6 and 6-<12 were associated with ‘ASD with ID’ (Figure S5).

### 3.4 CNS and non-CNS infections

Larger risk estimates for all outcomes were generally observed for CNS infections than for non-CNS infections (Figure 4), e.g. risk for ‘ID without ASD’ was 3.66 (95% CI: 2.91-4-60, figure 4E) for CNS infections and 1.42 (95% CI: 1.27-1.58, figure 4F) for non-CNS infections.

**Figure 4.**
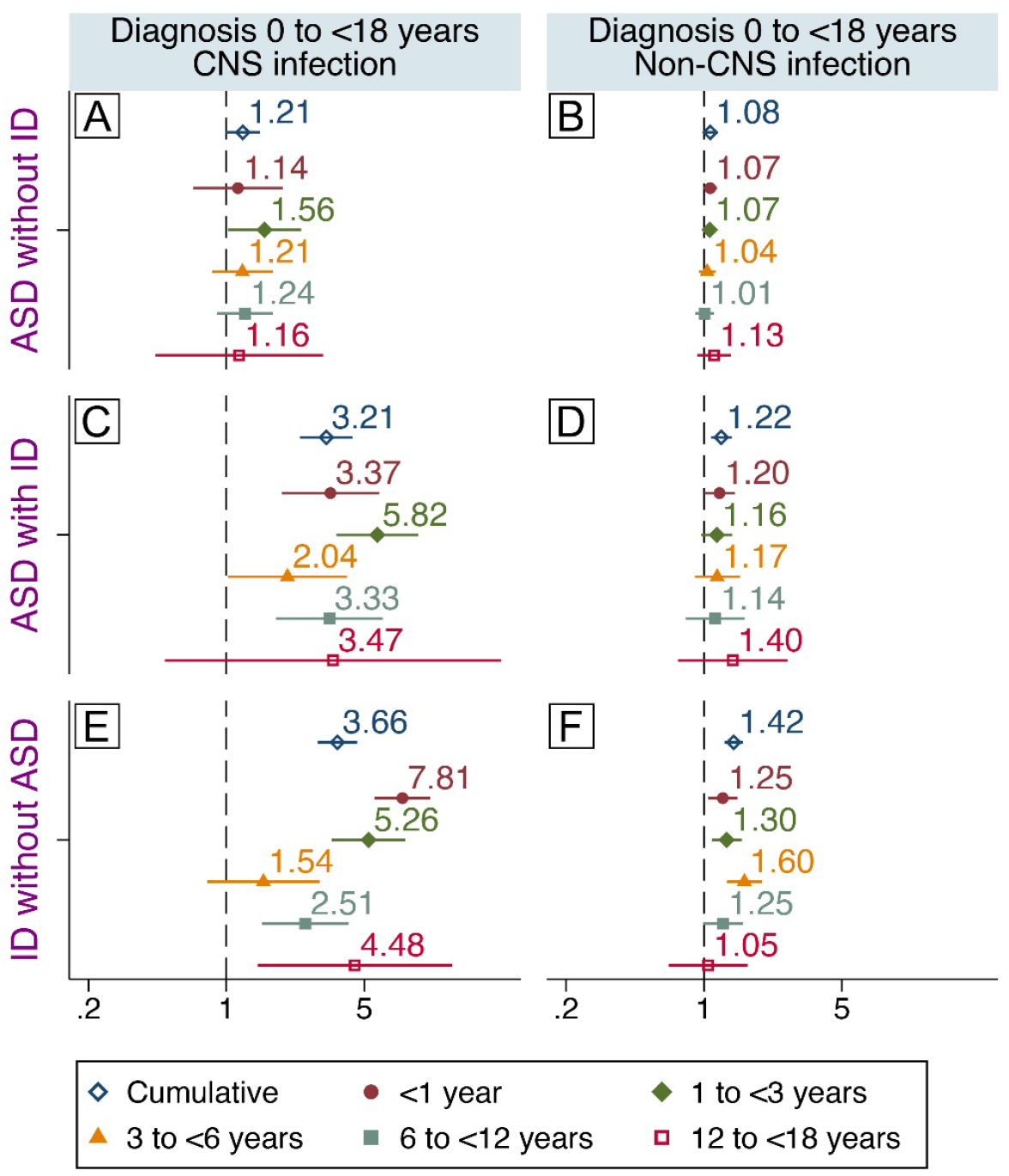
CNS/non-CNS infections and mutually exclusive diagnostic groups. Associations between specialized care for CNS infections (left) or non-CNS infections (right) during childhood and later diagnosis of ‘ASD without ID’ (A-B), ‘ASD with ID’ (C-D) or ‘ID without ASD’ (E-F). Associations between exposure at different age intervals and later diagnoses are also shown. Hazard ratios are adjusted for sex, parity, maternal body mass index, pre-eclampsia, parental age, education, income, region of origin, histories of psychiatric illness and infections, season of birth, gestational age at birth, size for gestational age, cesarean section and Apgar score.

## 4. Discussion

In the present study, infections requiring specialized care between ages 1 and 6 were associated with an excess risk of ‘ASD without ID’ before age 12. Infections before age 12 were associated with ‘ASD with ID’ diagnosed throughout childhood. Similar observations were made for ‘ID without ASD’ with even larger point estimates, particularly for infections during the first few years of childhood. We noted larger risks for all outcomes associated with CNS infections than for non-CNS infections, particularly for diagnoses involving ID. While our analyses highlighted the potential for confounding by both familial factors and factors not shared between family members, they also indicate that childhood infections cannot be excluded in the etiologies of ASD/ID, including those diagnosed years later.

### 4.1 Agreement with previous studies

#### 4.1.1 Infections and ASD

Long-term follow-up studies of individuals with congenital infections have reported autism-like behaviors ^37-39^. There are, however, few published studies of the potential association between infections overall and ASD in the general population, none of which considered the potential differential effect on ASD with or without co-occurring ID or the potential for confounding by factors shared between family members.

Rosen et al. reported no association between inpatient, outpatient, emergency and referral visits for infections registered within the Kaiser-Permanente health plan during the first two years of life and later ASD in a sample consisting of 500 cases and 2000 matched controls ^30^. Atladottir et al., on the other hand, reported associations between hospitalization for infections during childhood and later ASD (HR 1.38) in a Danish population in 2010 ^29^, similar to those observed here (HR 1.23). More recently, Sabourin and co-workers reported that individuals with ASD were more likely to have had an infection during the first three years of life than comparison individuals using a combination of neonatal records and care-giver interviews prone to bias ^31^. Our observations regarding CNS-infections agree with those reported by Pedersen et al. ^18^, who found a weak association between CNS-infections and ASD that disappeared after exclusion of individuals with co-occurring ID, although we lacked power to distinguish between effects of CNS and non-CNS infections.

#### 4.1.2 Infections and ID

With regard to the observed risk for ID, our results are in agreement with a number of clinical follow-up studies of neonates with congenital infections ^13^ and CNS-infections ^40^ of both viral and bacterial origin during both infancy and childhood ^14-16, 41-43^. In a study employing Danish population registers ^18^, the authors explored a wide range of mental disorders in relation to previous CNS infections and reported a significant association (HR 3.29) between CNS-infections and later ID, similar to those observed here for ‘ID without ASD’ (HR 3.66) and for ‘ASD with ID’ (HR 3.21).

Fewer studies have focused on cognitive function after childhood infections not involving the CNS. Kariuki *et al*., reported impairments in neurocognitive testing at age 7 in children who had been hospital treated for infections before age four ^44^. Moreover, the Danish register study by Atladottir *et al*. reported an association between infections during childhood and later ID ^29^ of a magnitude similar to that observed here. Our current findings are in line with previous studies where both CNS and non-CNS infections during early childhood were associated with lower scores on a mandatory cognitive test administered to conscripts ^21, 45^. Taken together with these previous studies, our current observations, suggest that infections without a recognized CNS involvement during childhood may contribute to impaired cognitive function.

### 4.2 Strengths and weaknesses

Strengths of the study include the large, population-based cohort with prospectively collected data in a setting with universal access to health care with minimal risk for selection or recall bias. Though all clinical pathways to ASD/ID diagnosis are covered, we do not have access to individual assessments of cognitive function. Thus, children diagnosed as ‘ASD without ID’ could experience impairments that do not meet criteria for ID. Despite the large study population, we had limited power to analyze specific infections.

Potential confounding was addressed in multiple ways. We adjusted our total-population estimates for wide range of potentially confounding factors. We used sibling comparison models to account for residual confounding by factors shared between siblings and adjusted for factors not necessarily shared between siblings (e.g. pre-term birth). Sensitivity analyses indicated that the associations observed were not fully explained by higher risk of infection among children with congenital diagnoses, although confounding by undiagnosed genetic abnormalities cannot be excluded.

Age at diagnosis depends on a wide range of factors such as clinical practice, severity and parental vigilance ^1^. Results from studies using register-based data to investigate timing of an exposure in relation to the onset of a childhood disorder should therefore be interpreted cautiously. While infections during early childhood were more common in children diagnosed with ASD or ID than in children without such diagnoses, we also note that the group diagnosed with ID at a young age had an excess of infections throughout childhood. Such an excess of infections in the group of individuals diagnosed with ASD without ID, however, seemed to be limited to the first decade of life. While these groups contain individuals with underlying hereditary and non-hereditary causes of ASD/ID that also receive more specialized care for infections during childhood (as addressed by our total population, sibling and sensitivity analyses), this does not preclude infections from being casually linked to some cases of ID and ASD. A higher liability to infections would increase the risk of contracting an infection requiring specialized care during early childhood, arguably a period of development more sensitive to external insults than later parts of childhood. It should also be kept in mind that our exposure definition is broad and encompasses a range of infectious agents, each with a prevalence and consequences that are likely to vary by age ^46^, which complicates comparisons of effects of exposures at different ages.

### 4.3 Potential mechanisms underlying the observed associations

A number of different, but not mutually exclusive, mechanisms can potentially explain our current observations. Targeting of the nervous system by infectious agents during periods of rapid growth and plasticity ^47^ is one plausible mechanism. Complications involving the CNS are, albeit rarely, observed for many different infectious agents, including those not normally associated with neuro-invasion, particularly in young individuals ^46, 48, 49^. For example, respiratory viruses (e.g. influenza A virus, respiratory syncytial virus) are important causes of encephalitis in young children ^50^. It is possible that for some registered infections, mild CNS-involvement may have gone unnoticed or undocumented, as has been reported for uncomplicated measles virus infections ^51^. Moreover, low levels of several molecules involved in the innate immune response at birth were recently associated with ASD risk ^24^, supporting the notion of an increased vulnerability to infections in ASD that was proposed based on a genetic correlation between hospitalization for infection and ASD ^52^. Further studies are needed to investigate if deficits in immunity associated with ASD risk could contribute to more severe outcomes of childhood infections, such as ID, and potentially contribute to the high co-morbidity between these disorders ^53^.

Direct support for immune mediated influences on the developing CNS resulting in cognitive and behavioral abnormalities is provided by experimental studies. Various protocols mimicking viral or bacterial infections during certain stages of development produce persistent cognitive deficits and behavioral changes in adult animals reminiscent of those observed in individuals with ASD ^54^. Moreover, transient treatment with interferon-γ during neuronal differentiation of human stem cells was reported to cause induction of ASD-associated genes ^55^. While studies of individuals diagnosed with ASD support activation of the immune system in ASD, the pathogenetic role of such immune activity remains unclear ^56-59^. Nevertheless, many of the pathways activated by infections have pleiotropic effects during brain development ^60^. Thus, the association between infections during periods of rapid growth and plasticity of the brain and later diagnosis of ASD/ID may be explained by components of the immune system interfering with the normal trajectories of brain development.

Infections cause changes in the distribution of both macro- and micronutrients in the host, with the purpose of promoting the function of the immune system while limiting pathogen proliferation ^61, 62^. Indeed, febrile episodes during childhood have been associated with reduced adult height, independent of genetic background and nutritional intake ^63^. Brain development during the first years of life is characterized by rapid, sequential patterns of cellular differentiation, migration and synapse formation during which the demand for macro- and micronutrients is high ^64^. Diversion of macro- and micronutrients necessary to mount an adequate immune response and clear an infection may explain part of our observed association between infections during childhood and later ID/ASD ^65, 66^.

### 4.4 Conclusions

Infections at an early age, appear to confer risk for a later diagnosis of ASD involving ID. Our observation that infections during early childhood exhibit weak but significant associations with ‘ASD without ID’ requires replication in future studies. Pending independent replication, future studies should focus on identifying specific infectious agents, their mechanisms of action, and individuals at risk. Such information will be critically important for guiding the development of preventive strategies.

## Data Availability

The Swedish health and population register data used in this study are available from Statistics Sweden and the Swedish National Board of Health and Welfare. The authors are not allowed to distribute the data according to the ethical approval for this study and the agreements with Statistics Sweden and the Swedish National Board of Health and Welfare.

## Acknowledgments

This work was generously supported by the Stanley Medical Research Institute (V-002, HK) and the Swedish Research Council, grant numbers 2018-02907 (HK), 2016-01477, 2012-2264 and 523-2010-1052 (CD)

## Supplementary material

**Supplementary figure S1.**
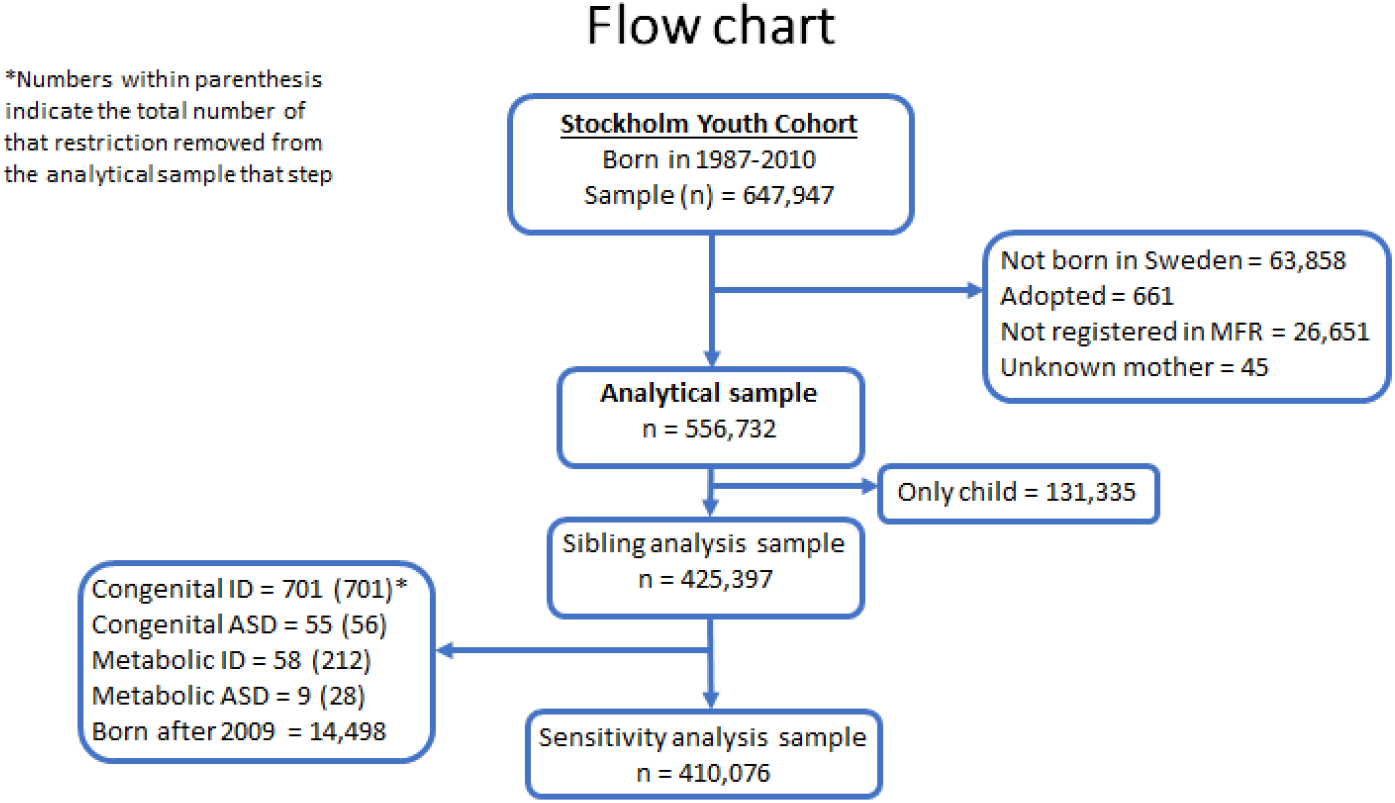
Flow chart delineating the derivation of the populations included in the present study.

**Supplementary figure S2.**
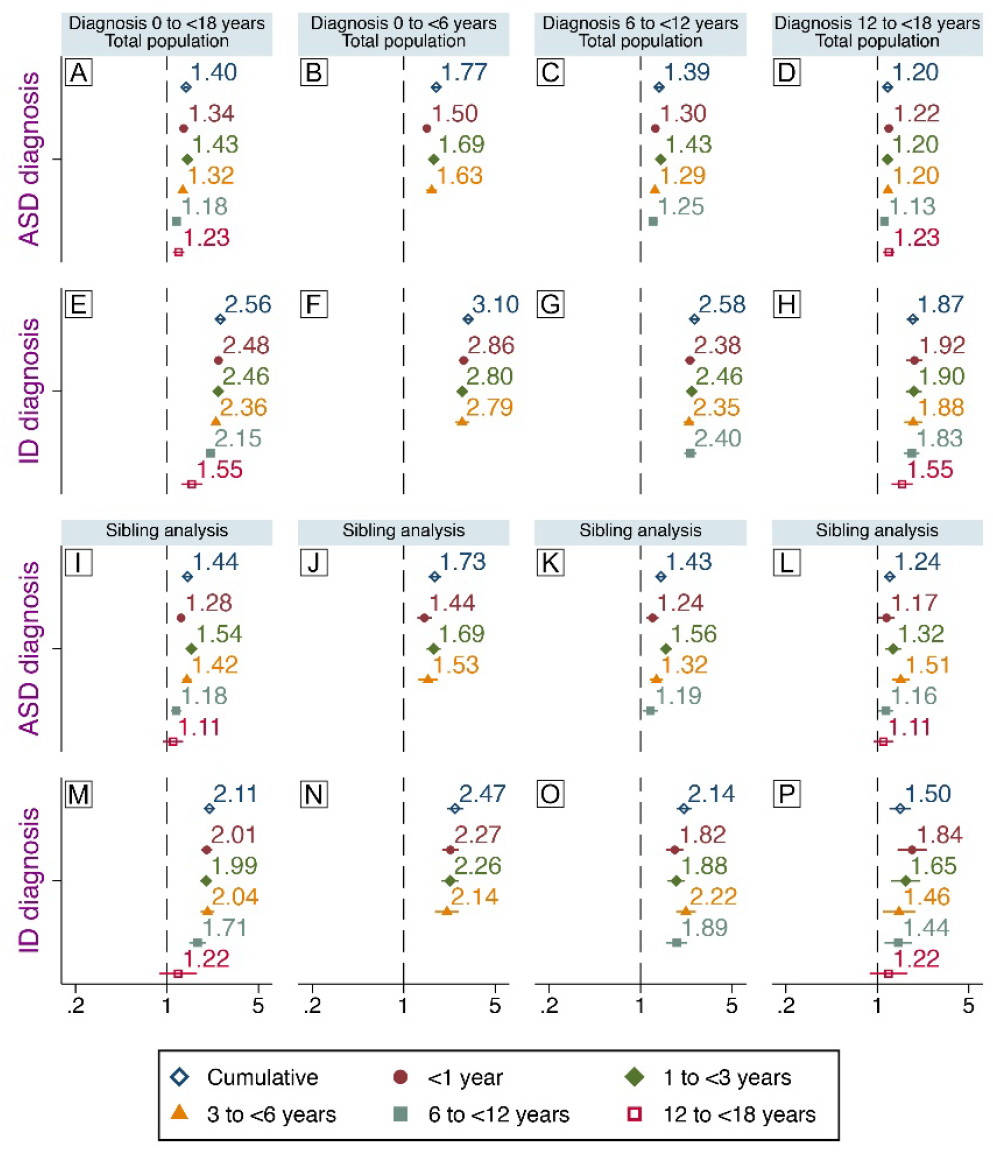
Infections during childhood and later diagnosis of ASD or ID. Crude associations between specialized care for infections and later, non-mutually exclusive, diagnosis of ASD and ID. Associations between exposure and diagnoses at different age interval are also shown. Comparison between unrelated individuals in the general population are shown (A-H) and comparisons between full biological siblings (I-P).

**Supplementary figure S3.**
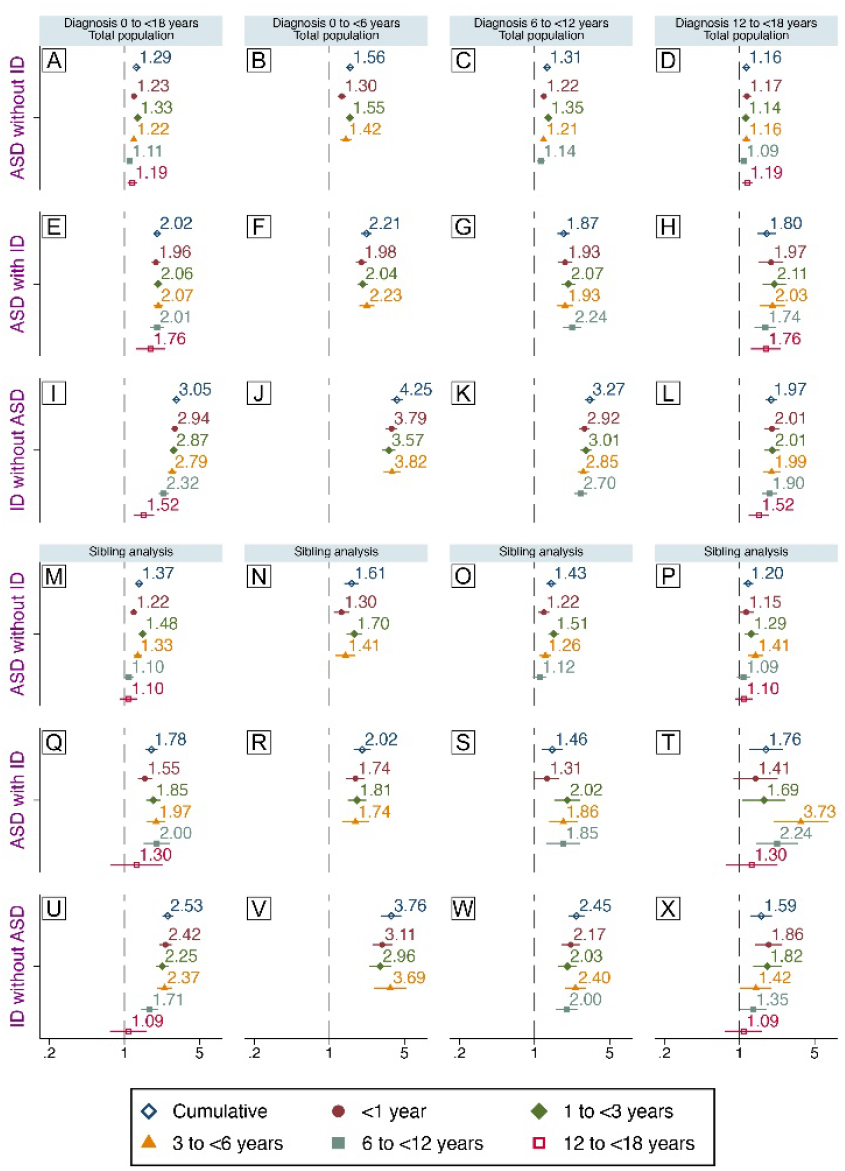
Infections during childhood and mutually exclusive diagnoses. Crude associations between infections between birth and age 18 and later diagnosis of ‘ASD without ID’, ‘ASD with ID’ or ‘ID without ASD’. Associations between exposures and diagnoses at different ages are also shown. Comparisons between unrelated individuals in the general population (A-L) and between full biological siblings (M-X) are shown.

**Supplementary figure S4.**
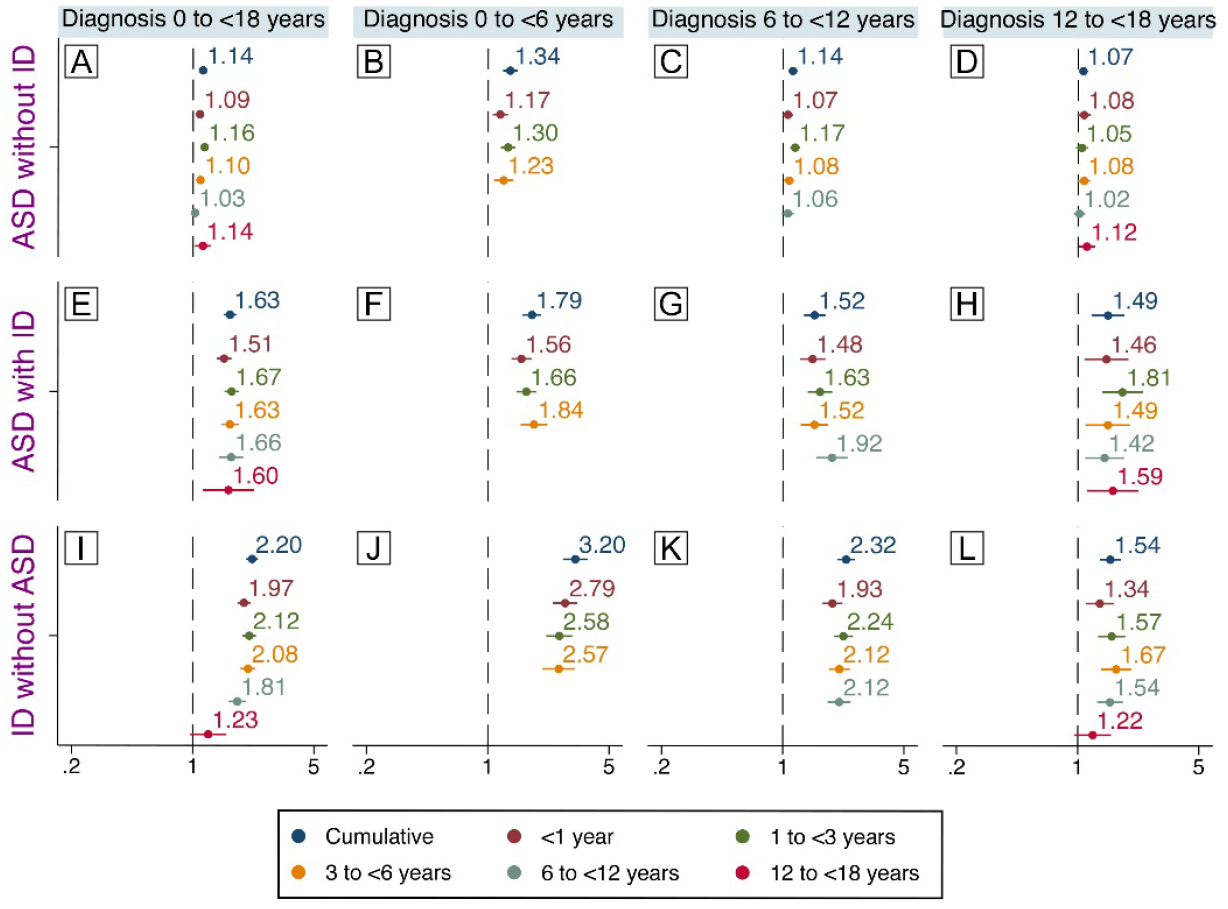
Infections during childhood and mutually exclusive diagnoses-sensitivity analyses. Associations between infections between birth and age 18 and later diagnosis of ‘ASD without ID’, ‘ASD with ID’ or ‘ID without ASD’ in the total population following exclusion of individuals with congenital diagnoses and ASD/ID. Associations between exposures and diagnoses at different ages are also shown. Estimates are adjusted for sex, parity, maternal body mass index, pre-eclampsia, parental age, education, income, region of origin, histories of psychiatric illness and infections, season of birth, gestational age at birth, size for gestational age, cesarean section and Apgar score.

**Supplementary figure S5.**
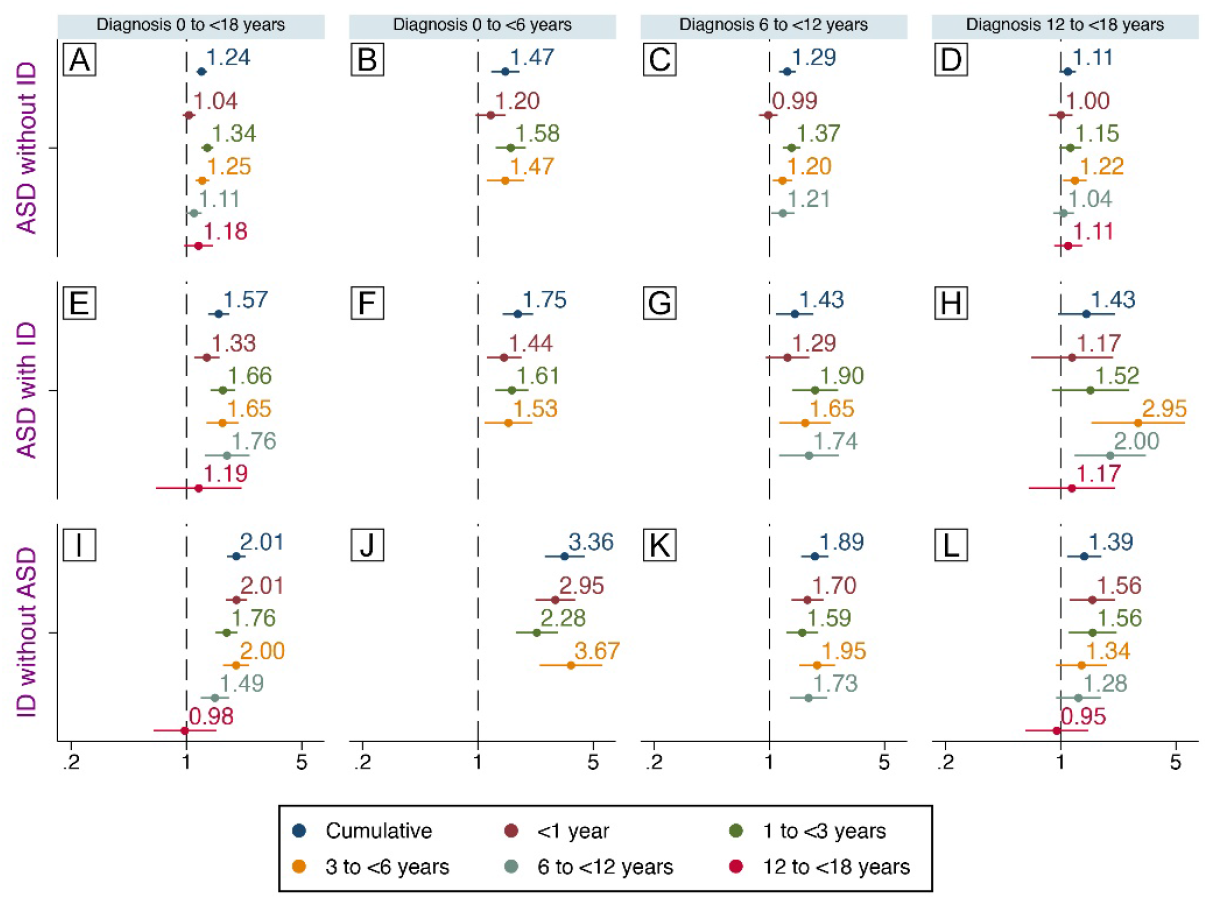
Infections during childhood and mutually exclusive diagnoses-sensitivity analyses. Associations between infections between birth and age 18 and later diagnosis of ‘ASD without ID’, ‘ASD with ID’ or ‘ID without ASD’ among biological siblings following exclusion of individuals with congenital diagnoses and ASD/ID. Associations between exposures and diagnoses at different ages are also shown. Estimates from are adjusted for sex, parity, gestational age at birth and cesarean section.

**Supplementary Table S1.**
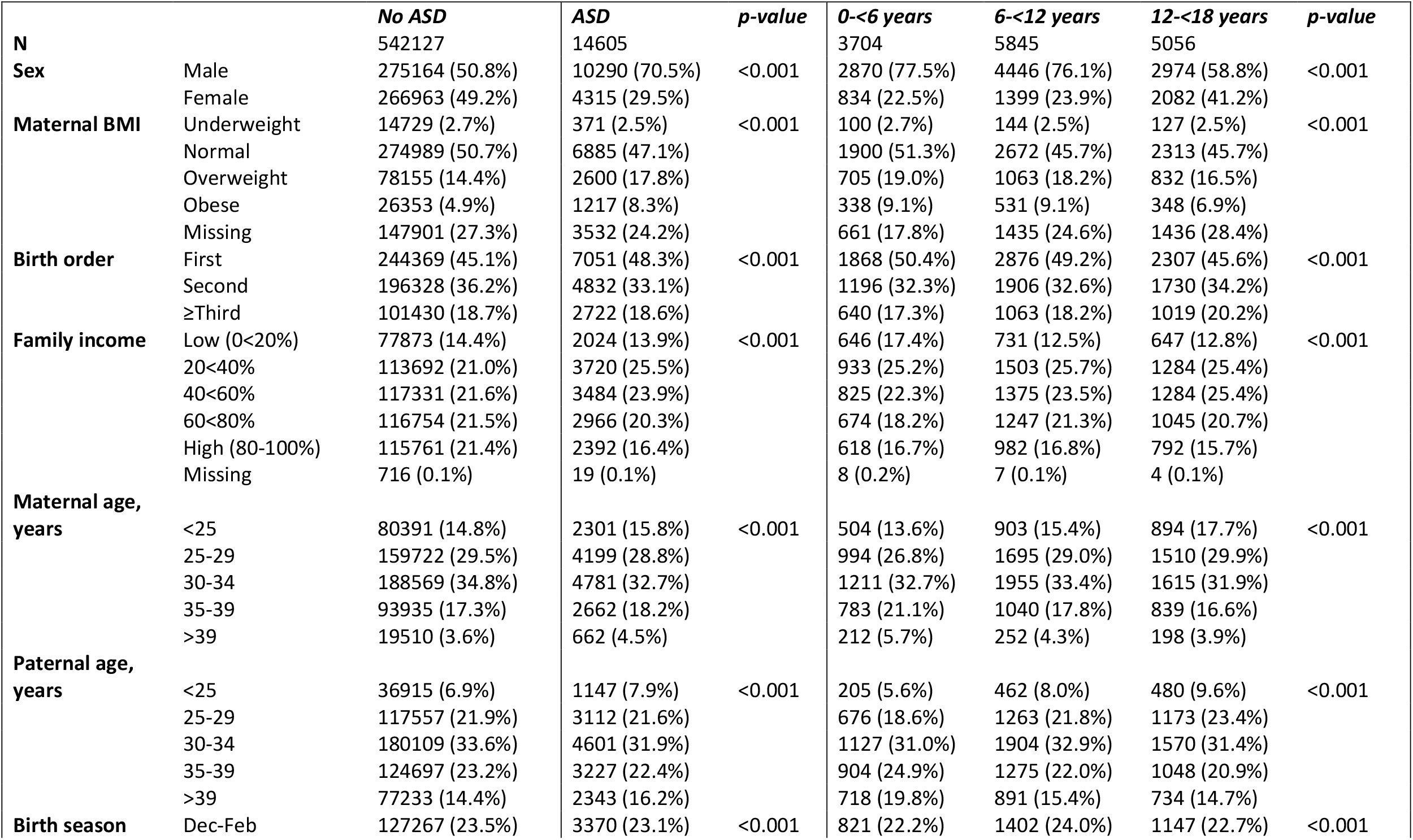
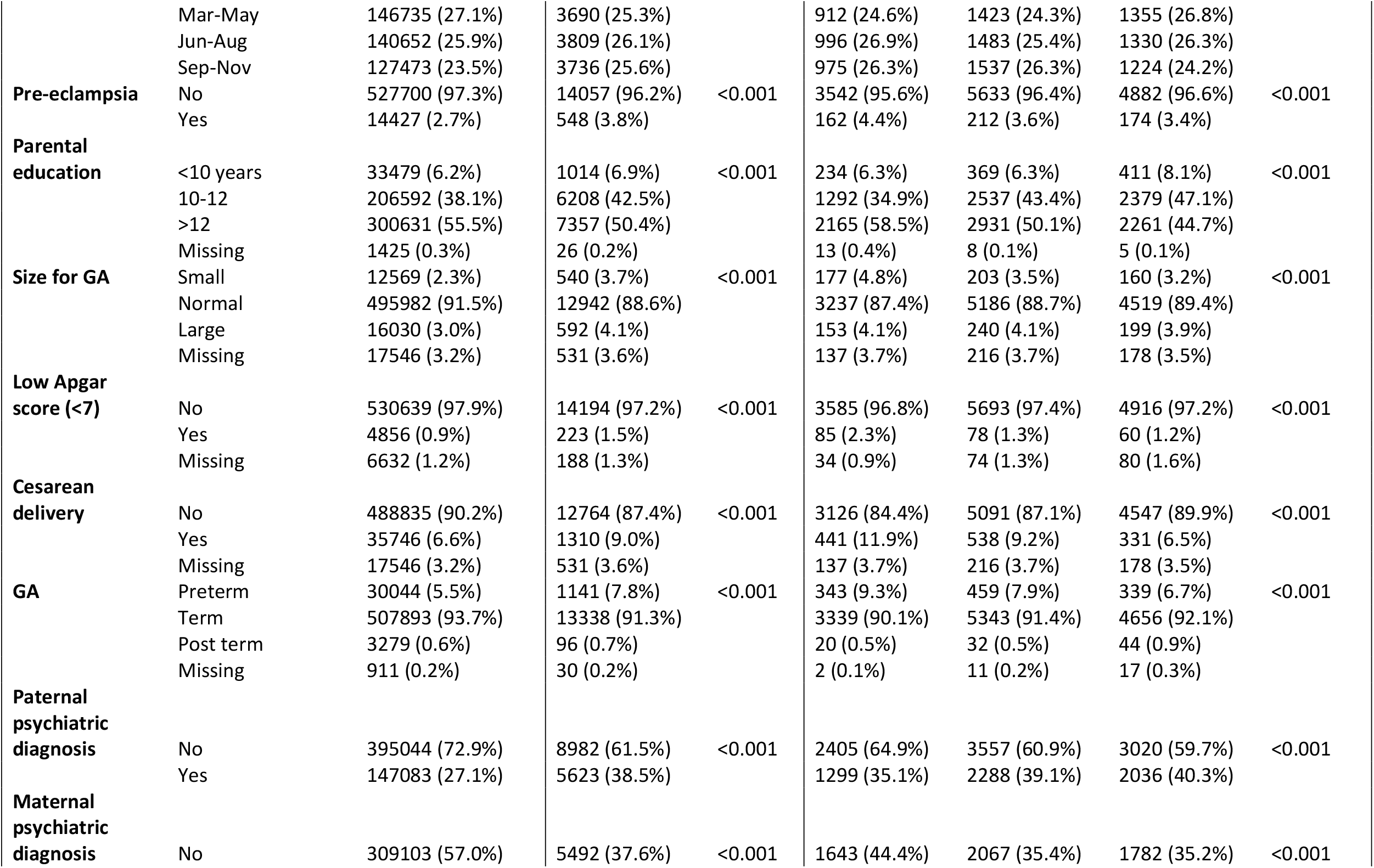
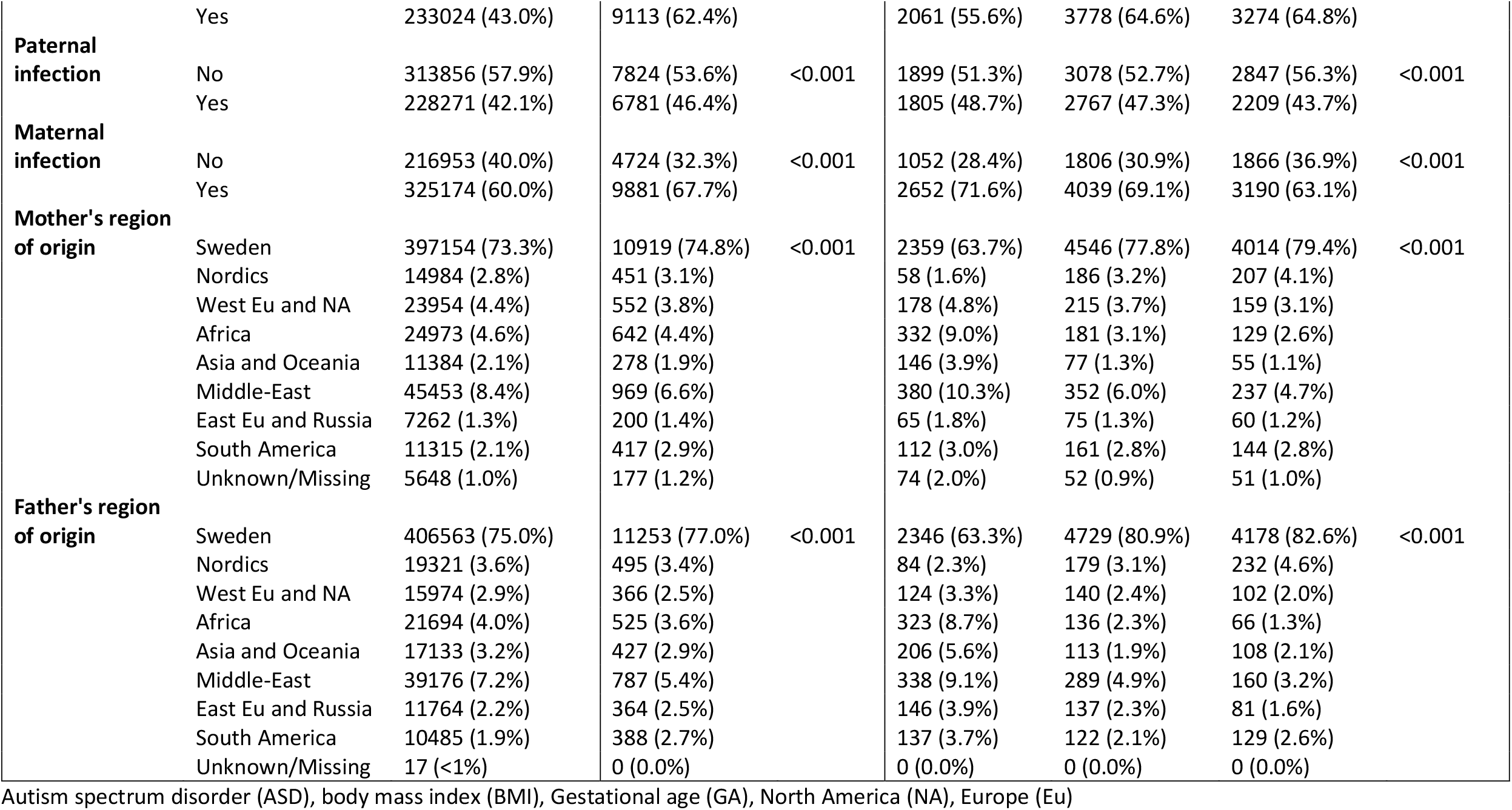
Association between covariates and ASD and by age at first diagnosis.

**Supplementary Table S2.**
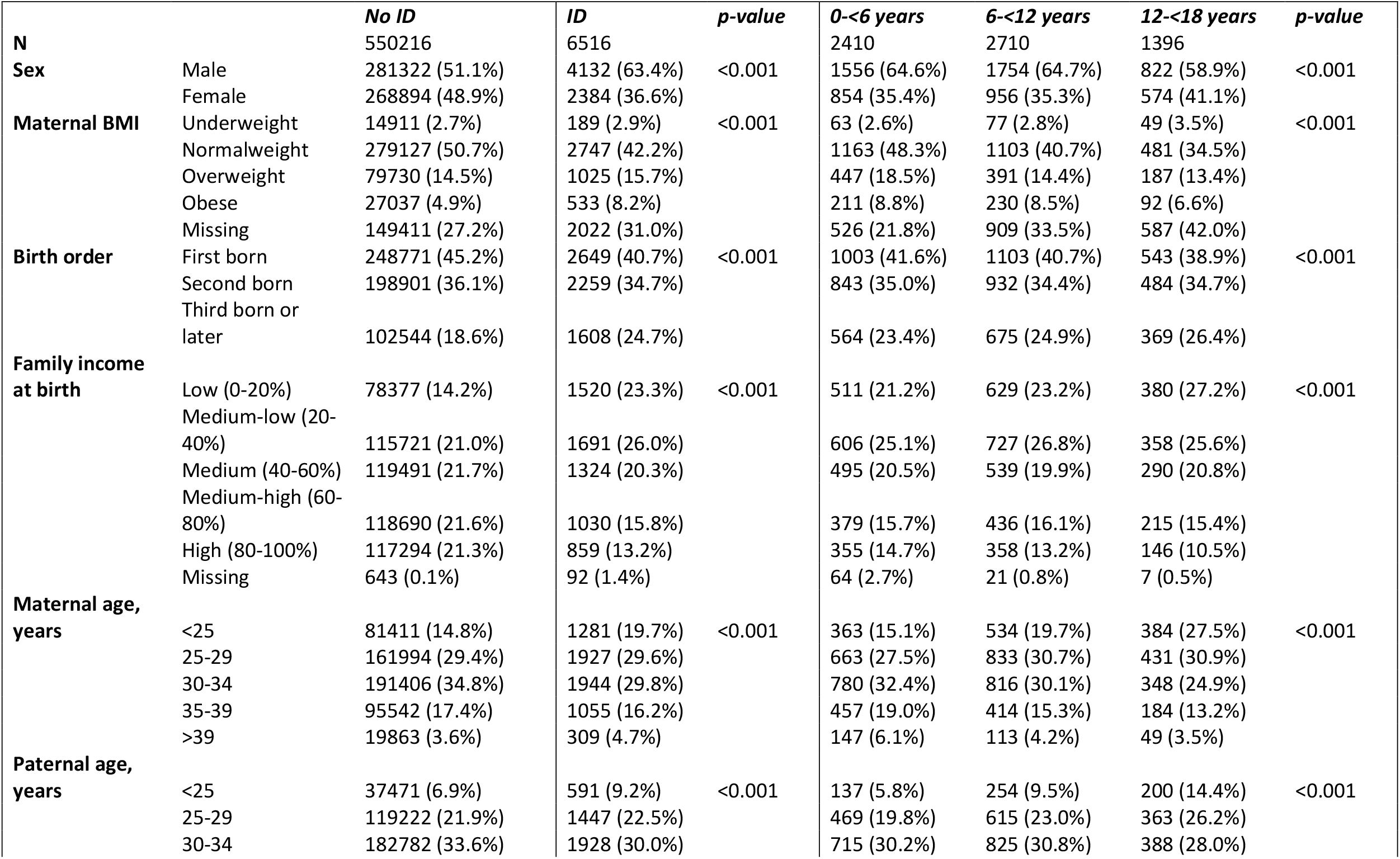
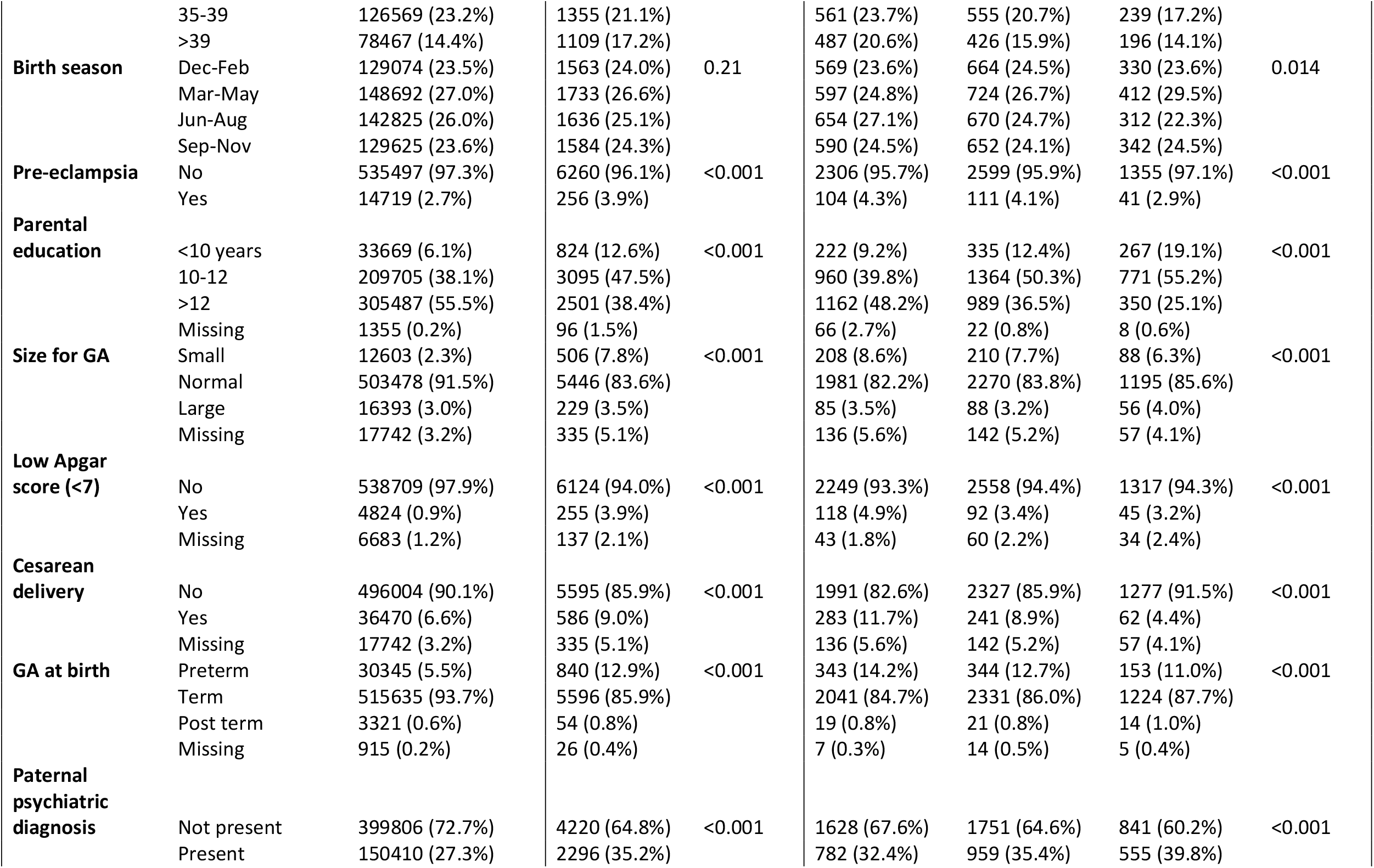
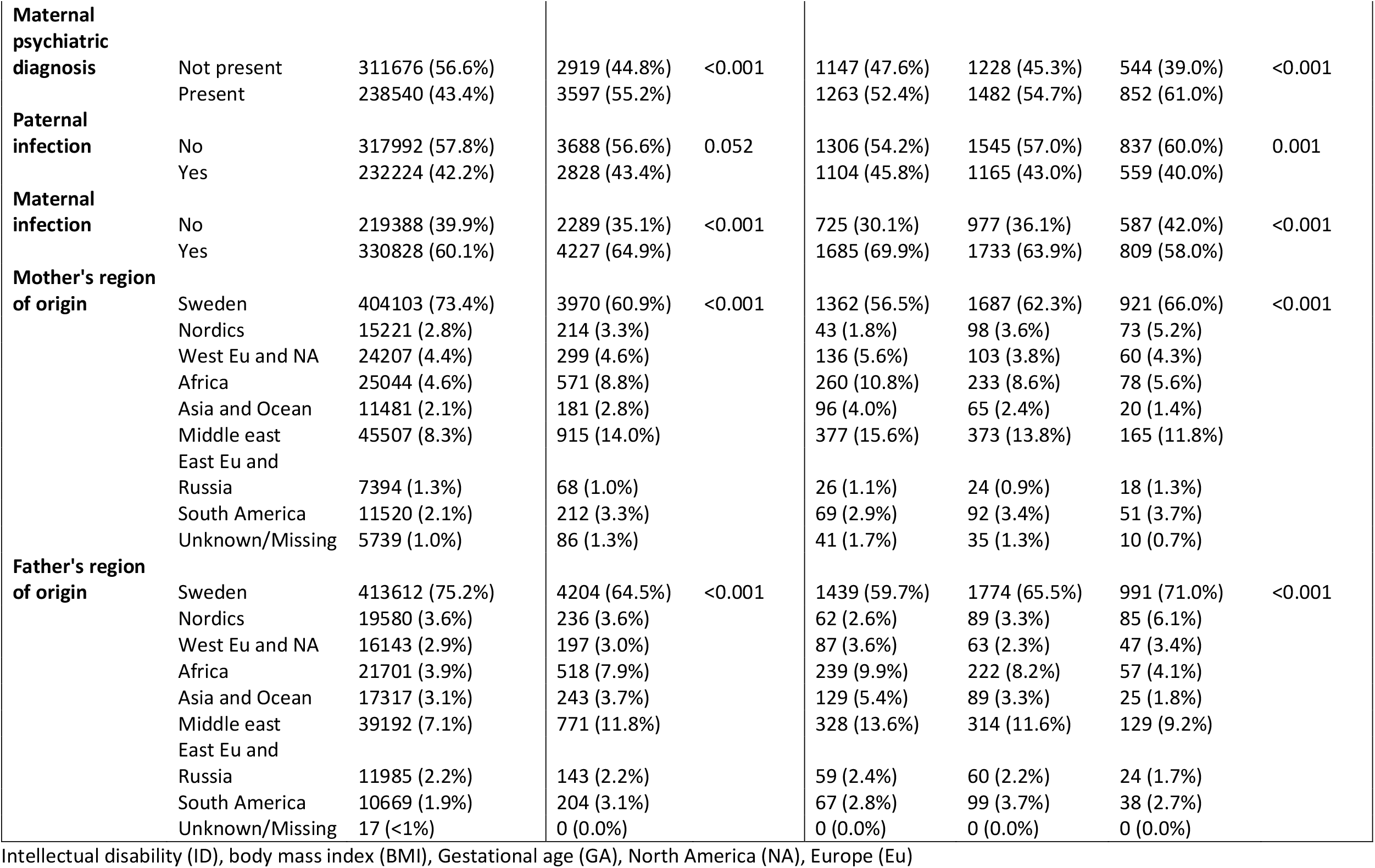
Association between covariates and ID and by age at first diagnosis.

## Notes

### Competing Interest Statement

The authors have declared no competing interest.

### Funding Statement

This work was generously supported by the Stanley Medical Research Institute (NV-002, HK) and the Swedish Research Council, grant numbers 2018-02907 (HK), 2016-01477, 2012-2264 and 523-2010-1052 (CD)

### Author Declarations

Stockholm Regional Ethics Committee, #2010/1185-31/5.

